# Gabapentinoid-to-Loop Diuretic Prescribing Cascade: A Systematic Review and Meta-Analysis of Observational Studies

**DOI:** 10.64898/2026.11.08.26367537

**Authors:** Abdallah Alami, Lucia de la Mora Vargas, Isabella Prada Betancourt, Lucía López-Paternostro, Emilio A. Flores Boggiano, João Victor M. S. Martelli, Lívian Scabeni Pan, Paulina Vivanco-Aravena, Oscar Munoz-Narvaez, Daniel Montanini, Marcela Borba Osorio, Sara Baroom, Rossy Cruz-Vicioso, Yuri Ramírez, Daniela Abele, Manuel Santos-Salcé, Carlos Collado, Isabela Resende da Glória, Fatima Ramos, Tatiane De Miranda, Lidiz Mora-Marquez, Abdul Haseeb, Federica Rossetto, Guilherme Franceschini Machado

## Abstract

**Introduction:** Gabapentinoid-induced peripheral edema may be misinterpreted as a new condition and treated with a loop diuretic, creating a prescribing cascade.

**Objective:** To synthesize evidence on the gabapentinoid-to-loop diuretic prescribing cascade in adults: its magnitude, dose- and drug-related gradients, timing, and downstream harms.

**Methods:** We searched MEDLINE, Embase, PubMed, and Web of Science to 23 April 2026 (PROSPERO CRD420261379968) for studies of loop diuretic initiation after gabapentinoid initiation and assessed risk of bias with ROBINS-I. Findings were synthesized by study design, and adjusted sequence ratios were pooled in a random-effects meta-analysis.

**Results:** Nine studies from five countries (>1.8 million new gabapentinoid users) were included: five sequence symmetry analyses, three cohort studies, and one chart review; six were restricted to older adults and two were available only as conference reports. All five sequence symmetry analyses and both cohort studies with an unexposed comparison group found more diuretic prescribing after gabapentinoid initiation. Sequence ratios ranged from 1.23 to 1.69, with a pooled adjusted sequence ratio of 1.37 (95% CI 1.14– 1.64; I^2^ = 96%; five estimates from four studies). Both of these cohort studies reported an adjusted hazard ratio of 1.44 (loop diuretics in one, any diuretic in the other). Higher doses were associated with larger estimates in four of the five studies that examined dose. In one chart review, 23.3% of patients had a potential harm within 60 days of starting a loop diuretic.

**Conclusions:** Gabapentinoid initiation was associated with subsequent loop diuretic prescribing, mainly in older adults, consistent with a prescribing cascade. Clinicians may consider a recently started gabapentinoid as a possible cause of new edema before adding a loop diuretic.

## Introduction

A prescribing cascade occurs when an adverse drug reaction is misinterpreted as a new medical condition and treated with a second medication, exposing the patient to additional drugs and risk while the cause goes unaddressed [1]. First described by Rochon and Gurwitz, prescribing cascades are now recognized as a form of inappropriate prescribing and avoidable drug-related harm, particularly among older adults [1, 2]. A recent review identified 84 distinct adverse drug reactions with the potential to trigger prescribing cascades [3].

Gabapentinoids (gabapentin, pregabalin, and mirogabalin) are among the most rapidly expanding drug classes. Their use among United States adults has risen markedly [4], and the estimated number of US emergency-department visits at which a gabapentinoid was prescribed at discharge rose from 138,479 in 2012 to 893,495 in 2021 [5]. Much of this growth reflects off-label prescribing for pain, despite limited evidence of efficacy for most such uses [6]. Peripheral edema is a recognized class effect and one of the most frequently reported adverse effects of gabapentinoids [7, 8]. It appears to be vasodilatory, resembling the edema caused by calcium channel blockers, and in pharmacovigilance data it often followed a dose escalation [9].

A loop diuretic may then be prescribed for new edema in a recent gabapentinoid user, either because the edema is misattributed to a cardiovascular or venous condition or as an avoidable symptomatic response, creating a gabapentinoid-to-loop diuretic prescribing cascade. This sequence mirrors cascades in which calcium channel blockers [10] and beta-blockers [11] are followed by loop diuretics. Since 2020, it has been examined in several countries and study designs [12, 13], but no systematic review has synthesized this evidence. To address this gap, we conducted a systematic review and meta-analysis to (i) characterize the temporal association between gabapentinoid initiation and subsequent loop diuretic prescribing, (ii) assess the magnitude, dose- and drug-related gradients, and consistency of the association across settings, and (iii) summarize the available evidence on downstream clinical harms following loop diuretic initiation.

## Methods

This systematic review and meta-analysis followed a protocol registered with PROSPERO (CRD420261379968) and is reported according to the PRISMA 2020 statement [14]. Screening, data extraction, and risk-of-bias assessment were performed independently by pairs of reviewers, with disagreements resolved by consensus or by a third reviewer.

### Eligibility criteria

We included studies of adults (≥18 years) who were new users of a gabapentinoid (gabapentin, pregabalin, or mirogabalin), defined by no gabapentinoid dispensing during a study-defined period before the first prescription, that evaluated loop diuretic initiation after gabapentinoid initiation. Eligible designs were prescription sequence symmetry analyses (which compare how often each of two drugs is started first among people who received both), cohort studies, and descriptive studies without a comparator (narrative synthesis only), using administrative claims, electronic health records, or dispensing data from outpatient, primary care, pharmacy, long-term care, or nursing-home settings. No restrictions were applied by language, country, or publication date. We excluded studies conducted only in people younger than 18 years, studies whose entire source population had a condition independently indicating loop diuretic therapy (e.g., heart failure or advanced chronic kidney disease), studies in which the gabapentinoid was not the initiating drug, and studies of gabapentinoid discontinuation alone.

### Data sources and search strategy

We searched MEDLINE (Ovid), Embase (Ovid), PubMed, and Web of Science Core Collection from inception to 23 April 2026, using subject headings and free-text terms for gabapentinoids, loop diuretics, prescribing cascades, peripheral edema, and sequence symmetry analysis (Supplementary Table S1), and screened the reference lists of included studies. Records were deduplicated in EndNote using a validated method [15] and then in Covidence before screening.

### Study selection and data extraction

Titles and abstracts, and then full texts, were screened in duplicate in Covidence. Two reviewers independently extracted data using a standardized form, including country, data source, setting, design, study period, population, exposure and definition of new use, comparator, outcome and time window, sample size, effect estimates with 95% CIs, adjustment factors, and subgroup and timing analyses.

### Risk of bias assessment

Risk of bias in studies with a comparison group was assessed with ROBINS-I (Risk of Bias in Non-randomized Studies of Interventions) [16] across seven domains. No validated risk-of-bias tool exists for sequence symmetry analyses; we therefore adapted ROBINS-I to these studies, focusing the confounding domain on factors that change over time, such as prescribing trends, because stable patient characteristics are controlled by the self-controlled design [17]. As prespecified, certainty of evidence was assessed narratively by risk of bias, consistency, precision, and directness, because no certainty-of-evidence tool has been designed for sequence symmetry analyses.

### Outcomes

The primary outcomes were the sequence ratio for loop diuretic initiation after gabapentinoid initiation (sequence symmetry analyses) and the incidence and relative risk of loop diuretic initiation among new gabapentinoid users (cohort studies). Secondary outcomes were time to loop diuretic initiation and downstream clinical harms over study-defined follow-up.

### Data synthesis

Findings were synthesized by design and effect measure. Adjusted sequence ratios (a value above 1 means the gabapentinoid was more often started first) from full-text sequence symmetry analyses with non-overlapping data sources were pooled in a random-effects meta-analysis (restricted maximum likelihood with the Hartung–Knapp– Sidik–Jonkman method [18]), and heterogeneity was summarized with the I^2^ statistic. Where too few studies were available for pooling, findings were synthesized narratively.

## Results

### Study selection and characteristics

The search identified 735 records; after removal of duplicates, 328 were screened and 11 full-text reports were assessed (Supplementary Figure S1). Nine studies were included [12, 13, 19–25]; one record was excluded for an ineligible design and another because it was an earlier conference report of an included study [21].

The studies were conducted in Canada, the United States, Denmark, France, and Japan and were published between 2020 and 2026 (Table 1). Five were prescription sequence symmetry analyses, three were retrospective cohort studies, and one was a chart review; two were available only as conference reports [24, 25], and for one of these the authors provided a poster with additional detail [25]. Among the six studies that reported the number of new users, it ranged from 7,867 to more than 1.5 million. Six studies were restricted to adults aged 60 years or older, and seven excluded patients with heart failure or chronic kidney disease. Most studies examined gabapentin and pregabalin; one examined pregabalin and mirogabalin, including inpatient prescriptions [22], and one pregabalin alone [23]. Populations partly overlapped between the two Veterans Affairs studies [19, 20] and between the two Danish data sources [13, 24].

**Table 1.**
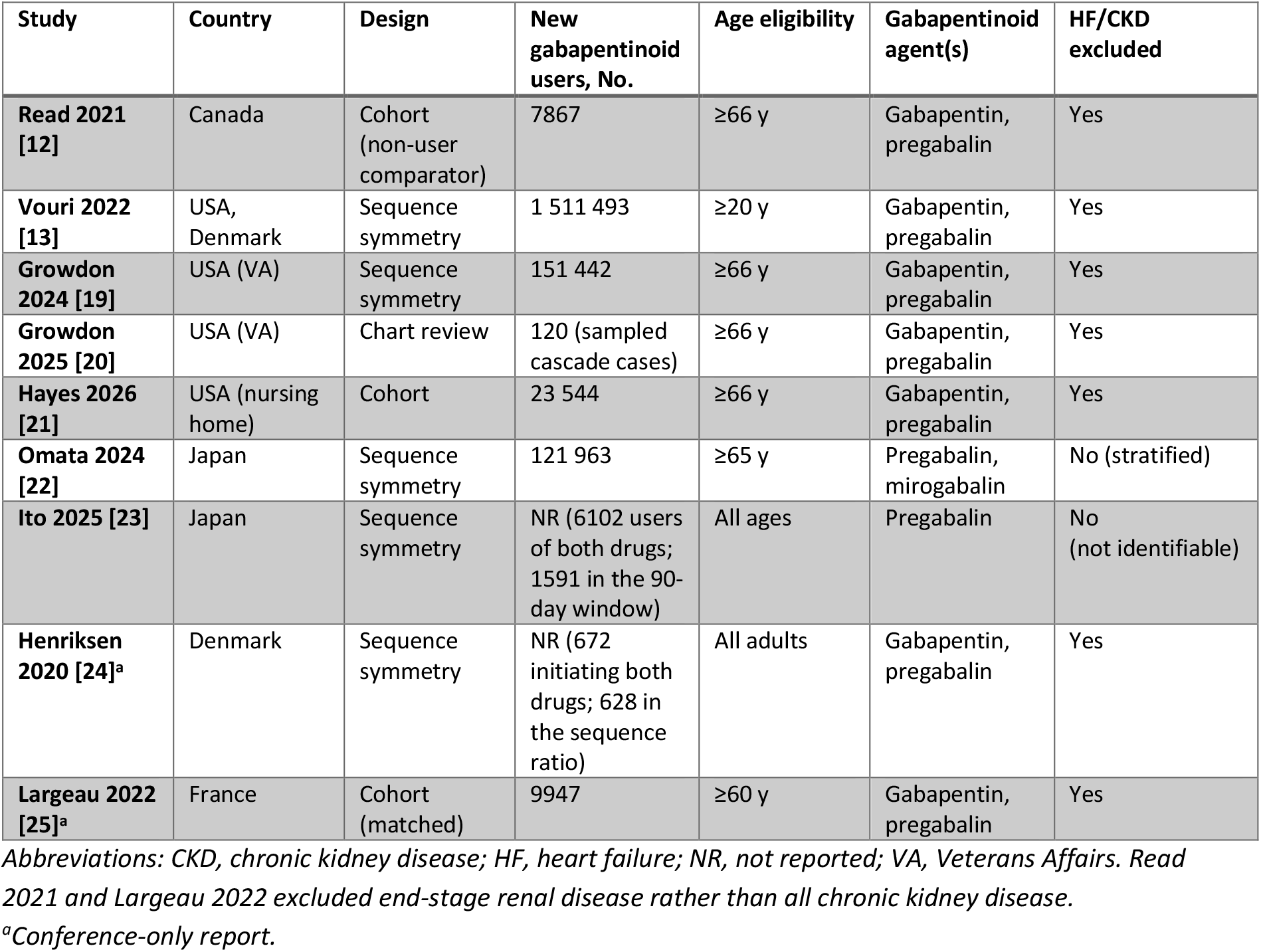
Characteristics of included studies.

| **Study** | **Country** | **Design** | **New gabapentinoid users, No.** | **Age eligibility** | **Gabapentinoid agent(s)** | **HF/CKD excluded** |
|---|---|---|---|---|---|---|
| **Read 2021 [12]** | Canada | Cohort (non-user comparator) | 7867 | ≥66 y | Gabapentin, pregabalin | Yes |
| **Vouri 2022 [13]** | USA, Denmark | Sequence symmetry | 1 511 493 | ≥20 y | Gabapentin, pregabalin | Yes |
| **Growdon 2024 [19]** | USA (VA) | Sequence symmetry | 151 442 | ≥66 y | Gabapentin, pregabalin | Yes |
| **Growdon 2025 [20]** | USA (VA) | Chart review | 120 (sampled cascade cases) | ≥66 y | Gabapentin, pregabalin | Yes |
| **Hayes 2026 [21]** | USA (nursing home) | Cohort | 23 544 | ≥66 y | Gabapentin, pregabalin | Yes |
| **Omata 2024 [22]** | Japan | Sequence symmetry | 121 963 | ≥65 y | Pregabalin, mirogabalin | No (stratified) |
| **Ito 2025 [23]** | Japan | Sequence symmetry | NR (6102 users of both drugs; 1591 in the 90-day window) | All ages | Pregabalin | No (not identifiable) |
| **Henriksen 2020 [24]**<sup>a</sup> | Denmark | Sequence symmetry | NR (672 initiating both drugs; 628 in the sequence ratio) | All adults | Gabapentin, pregabalin | Yes |
| **Largeau 2022 [25]**<sup>a</sup> | France | Cohort (matched) | 9947 | ≥60 y | Gabapentin, pregabalin | Yes |

*Abbreviations: CKD, chronic kidney disease; HF, heart failure; NR, not reported; VA, Veterans Affairs. Read 2021 and Largeau 2022 excluded end-stage renal disease rather than all chronic kidney disease.*
<sup>a</sup>*Conference-only report.*

### Magnitude and consistency of the association

All seven studies with a comparison group found more diuretic prescribing after gabapentinoid initiation, regardless of country, data source, follow-up window, or gabapentinoid examined (Table 2; Supplementary Table S2). Principal sequence ratios ranged from 1.23 to 1.69 [13, 19, 22–24]; pooling five estimates from four full-text studies gave an adjusted sequence ratio of 1.37 (95% CI 1.14–1.64), with high heterogeneity (I^2^ = 96%; Supplementary Figure S2). Within one study, the association was weaker in the United States (1.19; 95% CI 1.15–1.23) than in Denmark (1.50; 95% CI 1.43–1.57) [13]. The two cohort studies with an unexposed comparison group each reported an adjusted hazard ratio of 1.44, for any diuretic (95% CI 1.23–1.70) [12] and for loop diuretics (95% CI 1.20–1.73) [25]. Three studies reported absolute measures. Vouri et al. estimated about 8 excess loop diuretic starts per 1000 person-years of gabapentinoid use (8.1; 95% CI 1.9–34.5) [13], and Growdon et al. estimated 4.8 among US veterans [19]. Read et al. reported the difference as a proportion: within 90 days, 2.0% of new gabapentinoid users and 1.3% of non-users received any diuretic [12].

**Table 2.** Primary estimate of the gabapentinoid-to-loop diuretic cascade, by study.

| **Study** | **Measure** | **Point estimate (95% CI)** | **Time window** | **Outcome** |
|---|---|---|---|---|
| **Vouri 2022 [13]** | Adjusted sequence ratio | 1.33 (1.06–1.67) pooled;<br>US 1.19 (1.15–1.23);<br>Denmark 1.50 (1.43–1.57) | 90 d | Loop diuretic |
| **Growdon 2024 [19]** | Adjusted sequence ratio | 1.23 (1.13–1.34) | 180 d | Loop diuretic |
| **Omata 2024 [22]** | Adjusted sequence ratio | 1.69 (1.56–1.83) | 90 d | Loop diuretic |
| **Ito 2025 [23]** | Adjusted sequence ratio | 1.29 (1.16–1.43) | 90 d | Loop diuretic |
| **Henriksen 2020 [24]<sup>a</sup>** | Sequence ratio | 1.56 (1.34–1.84) | 90 d | Loop diuretic |
| **Read 2021 [12]** | Adjusted hazard ratio | 1.44 (1.23–1.70) | 90 d | Any diuretic |
| **Largeau 2022 [25]<sup>a</sup>** | Adjusted hazard ratio | 1.44 (1.20–1.73) | 120 d | Loop diuretic |
| **Hayes 2026 [21]** | Cumulative incidence; dose-response RR | 4.2% (994/23,544);<br>higher- vs lower-dose RR 1.45 | 90 d | Loop diuretic |
| **Growdon 2025 [20]** | Harm frequency | 23.3% with any potential downstream event | 60 d | Downstream harm |

*Read 2021 measured any diuretic initiation; furosemide accounted for 59.0% of diuretic prescriptions in the gabapentinoid group and 46.9% in the comparison group. Growdon 2025 reported descriptive harm frequencies without a comparator group. Hayes 2026 had no unexposed comparator. For Ito 2025, the 90-day estimate is shown to match the other studies' windows. The measures differ and are not directly comparable. <sup>a</sup>Conference only report.*

Two studies included a negative-control drug class, serotonin–norepinephrine reuptake inhibitors (SNRIs), which share some indications with gabapentinoids but are not commonly associated with edema. Neither showed excess loop diuretic prescribing after SNRI initiation [13, 23], and in one study the gabapentinoid sequence ratio was 1.58 times that of the negative control [13].

### Dose- and drug-related gradients

Of the five studies that examined dose, four found larger associations at higher doses, two of them only in secondary analyses or analyses of dose during follow-up [12, 21], and one found no difference by dose [25] (Figure 1; Supplementary Table S2). Sequence ratios were higher with doses at or above versus below the defined daily dose (1.52 vs 1.17) [13] and with a pregabalin starting dose of at least 150 mg/day than with a lower dose (1.53 vs 1.19; P = .02) [23]. In the cohorts, high-dose exposure during follow-up was associated with diuretic dispensing at 180 days (adjusted hazard ratio 2.34; 95% CI 1.17–4.67) [12], and a higher weekly dose during follow-up with a higher cumulative incidence of loop diuretic initiation (7.2% vs 5.0%; risk ratio 1.45), whereas baseline dose was not statistically significant after adjustment (adjusted risk ratio 1.21; 95% CI 0.99–1.48) [21]. The sequence ratio was larger for mirogabalin than for pregabalin (2.23 vs 1.59) [22], a between-drug comparison that may reflect differences in the patients receiving each agent. Because dose definitions differed and dose was not randomized, these gradients may partly reflect severity, treatment duration, or patient characteristics.

**Figure 1.**
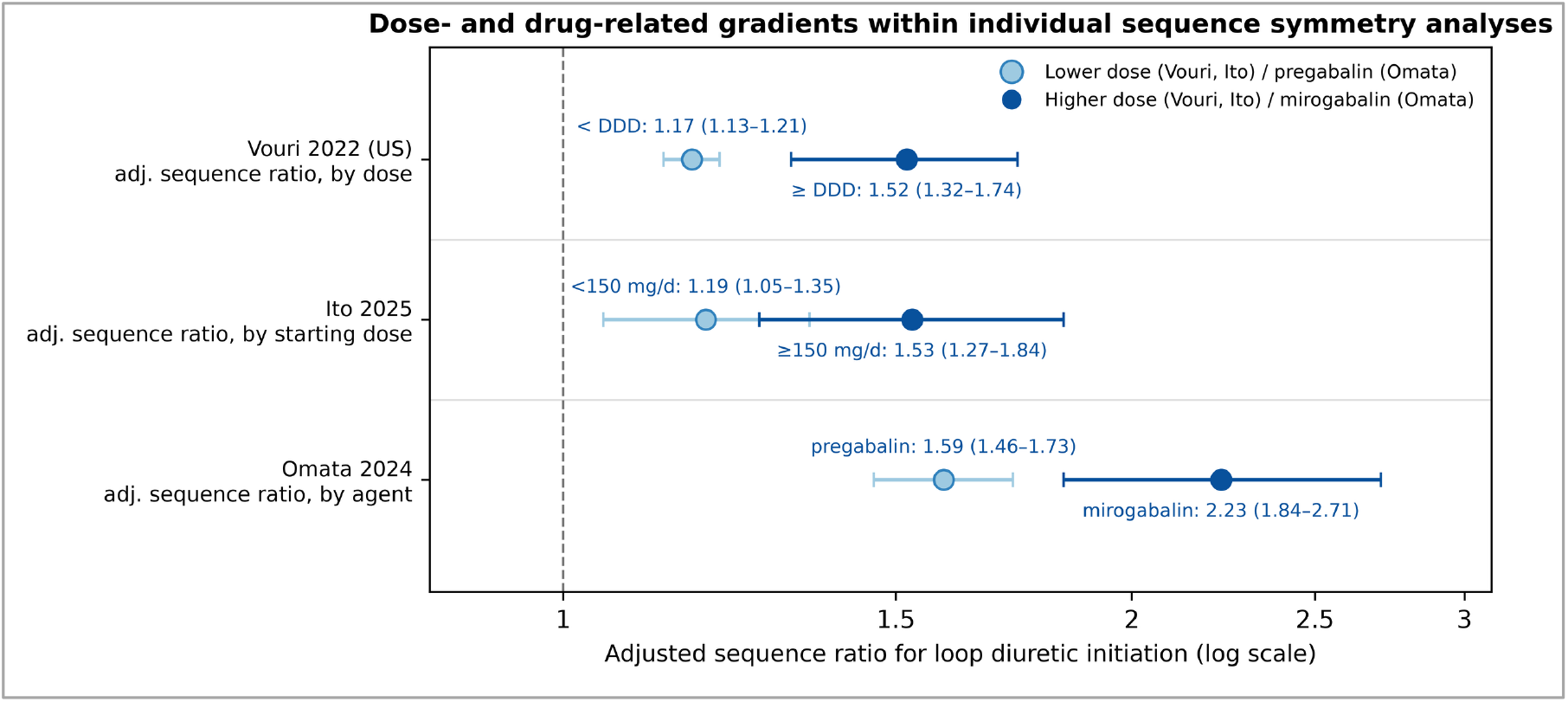
Dose- and drug-related gradients in adjusted sequence ratios within individual studies. Each row is a separate within-study comparison with its own population, window, and dose definition; rows share an axis for reference only and must not be read as a pooled or continuous dose-response gradient. DDD, defined daily dose.

### Timing of the cascade

Excess loop diuretic initiation was generally concentrated in the first months after gabapentinoid initiation. The median time to diuretic initiation was 28 to 36 days [12, 21], and sequence ratios were generally largest in the shortest observation windows [22, 23], although one cohort found no excess in the first 15 days [25].

### Subgroups and predictors

Within the sequence symmetry analyses, the association was present in most subgroups without consistent differences, including in patients with and without heart failure (1.62 and 1.84) [22]; estimates by age were broadly similar in the three studies that included younger adults [13, 23, 24], and no subgroup reversed the direction of the association (Supplementary Table S2). Predictors among new users pointed in different directions: in nursing homes, risk was higher at ages 86–90 and ≥91 years (adjusted risk ratios 1.60 and 1.38 vs 66–70 years) and lower with Alzheimer disease and related dementias (0.79) [21], whereas veterans aged 85 years or older were less likely than those aged 66–74 years to have the cascade sequence (adjusted odds ratio 0.74; 95% CI 0.56–0.96) [19].

### Downstream harms

Only one study examined events after loop diuretic initiation: a chart review of 120 veterans with documented edema, randomly sampled from a national cohort [20]. Within 60 days, 28 patients (23.3%) had 37 potential events, 10 of which were judged probably attributable to the loop diuretic; the most frequent were worsening kidney function (7.5%), orthostasis (5.8%), and electrolyte abnormalities (5.0%), and 4.2% had a fall and 5.0% an emergency visit or hospital admission [20]. Clinicians named a gabapentinoid in the differential diagnosis for edema in only 3.3% of cases but cited congestive heart failure, a baseline exclusion criterion, in 39.2% [20].

### Risk of bias and certainty of evidence

Of the seven studies with a comparison group, none was at low risk of bias: three were at moderate and four at serious risk (Supplementary Figure S3). The two descriptive studies without a comparison group were not assessed with ROBINS-I [20, 21]. The main concerns were the handling of conditions that themselves call for a loop diuretic, such as heart failure, in the sequence symmetry analyses, and baseline differences between users and non-users in the cohort studies; both designs rely on dispensing as a proxy for treated edema. Handling of other drugs that can cause edema varied across studies (Supplementary Table S3). Overall, confidence was greatest for the direction of the association, which was consistent across seven studies, five countries, and both main designs, but was limited by risk of bias. Confidence was lower for dose- and drug-related gradients, which used heterogeneous definitions, and lowest for downstream harms, which rest on a single uncontrolled chart review.

## Discussion

In this systematic review and meta-analysis of nine studies from five countries, including more than 1.8 million new gabapentinoid users, gabapentinoid initiation was consistently followed by excess diuretic prescribing. In sequence symmetry analyses, loop diuretics were more often started after than before a gabapentinoid (pooled adjusted sequence ratio 1.37), and in cohort studies, new users were more likely than non-users to start a diuretic (adjusted hazard ratio 1.44). The consistency of the association across countries, data sources, designs, and gabapentinoid agents, together with negative-control comparisons, dose-associated gradients, and a plausible temporal pattern, is consistent with a prescribing cascade. Because the evidence is observational and most studies could not confirm edema or the diuretic indication, it describes a potential rather than a clinically confirmed cascade. Given the rapidly expanding use of gabapentinoids [26], even a modest absolute excess could translate into a considerable population burden.

Several features support this interpretation, although none establishes causality. Sequence symmetry analyses, in which each person serves as their own control, and cohort studies with unexposed comparison groups are susceptible to partly different biases, so their agreement is reassuring; however, observational studies can share biases, such as detection bias and time-varying confounding. Comparison with SNRIs, which share indications with gabapentinoids but are not commonly associated with edema, reduces but does not eliminate concern about confounding by indication [13, 23]. Higher doses were associated with larger estimates in most studies that examined dose, but dose was not randomized and may reflect severity, treatment duration, or tolerance of the drug; we therefore interpret these findings as dose-associated gradients rather than a consistent dose-response relationship.

The timing is also coherent with the pharmacology of gabapentinoid-associated edema, which in adverse-event reports had a median onset of 23 days and most often followed a dose escalation [9]. Sequence ratios declined as observation windows lengthened, consistent with dilution by background prescribing over time, whereas one cohort found an excess only after the first 15 days [25]. This difference may reflect the different measures or delays between edema onset, recognition, and prescribing, and cannot be resolved from a conference report.

The magnitude of the association varied across countries, possibly reflecting differences in prescribing practices, access to specialist care, formularies, or data capture. Retaining patients with heart failure or chronic kidney disease raises background loop diuretic use and may dilute the sequence ratio [22]. This variation, reflected in the very high heterogeneity of the meta-analysis, argues for reporting country- and setting-specific estimates rather than relying on a single summary estimate.

Evidence on the consequences of this cascade is limited to one small uncontrolled chart review, in which nearly one quarter of patients had a potential harm within 60 days of starting a loop diuretic [20]. Diuretic use has been associated with electrolyte disorders, syncope, and falls, particularly in older patients (shown, for example, for thiazide diuretics [27]). In the same chart review, clinicians rarely considered a gabapentinoid as a cause of edema, and in 60% of cases the loop diuretic was prescribed by a different clinician from the one who prescribed the gabapentinoid [20]; such prescriber discordance was also more common in potential calcium channel blocker–loop diuretic cascades than in a control drug pair [28]. Although the included studies did not compare management strategies, clinicians may reasonably consider whether a recently started gabapentinoid is contributing to new edema, after evaluating alternative causes, and whether dose reduction, discontinuation, or an alternative treatment is appropriate before adding a loop diuretic. Prescriber education, decision support, and pharmacy-based medication reconciliation could help to flag this sequence, although their effectiveness has not been evaluated. Because gabapentinoid use is growing [26] and may continue long term (beyond 90 days in more than one-fifth of older adults newly prescribed gabapentin after surgery [29]), the population at risk is likely to expand.

### Strengths and limitations

To our knowledge, this is the first systematic review of the gabapentinoid-to-loop diuretic prescribing cascade. It combined complementary study designs from five countries, covering more than 1.8 million new gabapentinoid users, and pooled sequence symmetry analyses restricted to full-text studies with non-overlapping data sources.

This review also has limitations. First, all included studies were observational, and eight of nine used administrative or claims data that could not confirm edema, the indication for the loop diuretic, or the indication for the gabapentinoid; the one study that reviewed clinical records included only patients with documented edema, confirming that the pathway occurs but not how often it explains excess prescribing [20]. Second, only the sequence symmetry analyses could be pooled, and heterogeneity was very high, so the pooled ratio summarizes these studies rather than estimating a single common effect. Third, two studies were available only as conference reports, which provide limited methodological detail. Fourth, all studies came from high-income countries and most were restricted to older adults, so the findings apply most directly to older adults, although estimates by age in the three studies that included younger adults were broadly similar.

Future studies should link prescribing data to clinical records to confirm edema and the diuretic indication; compare new gabapentinoid users with new users of a similar drug to address confounding; and compare strategies for managing gabapentinoid-associated edema (dose reduction or discontinuation versus diuretic treatment) and downstream harms, ideally in pragmatic trials.

## Conclusion

Gabapentinoid initiation was consistently associated with subsequent loop diuretic prescribing, mainly in older adults, a pattern consistent with a prescribing cascade, although the evidence is observational and heterogeneous. When new peripheral edema follows the start of a gabapentinoid, considering the drug as a possible cause, after evaluating alternative causes, may avoid unnecessary diuretic treatment, although the benefit of this approach requires comparative evaluation.

## Supporting information

PRISMA Checklist

## Declarations

### Ethics approval

Not required. This systematic review analyzed only aggregate data from published studies and conference reports; no individual participant data were collected.

### Reporting guideline

This review is reported according to the PRISMA 2020 statement; the completed PRISMA 2020 checklist is provided as a supplementary file.

### Data availability

All data analyzed in this review derive from the published reports of the included studies. Extraction templates and the data supporting the synthesis are available from the corresponding author on reasonable request.

### Use of artificial intelligence

A large language model (Claude, Anthropic) assisted with language editing and drafting revisions; all outputs were verified and edited by the authors, who take full responsibility for the content.

