## Supplementary Material for "Gabapentinoid-to-Loop Diuretic Prescribing Cascade: A Systematic Review and Meta-Analysis of Observational Studies"

### Supplementary Table S1. Full search strategies by database (inception to 23 April 2026)

| Database (platform) | Date searched | Records retrieved | Search strategy |
|---|---|---|---|
| **MEDLINE (Ovid)** | 23 April 2026 | 218 | 1 gabapentin.mp. or Gabapentin/<br>2 pregabalin.mp. or Pregabalin/<br>3 mirogabalin.mp.<br>4 gabapentinoid*.mp.<br>5 (neurontin or lyrica or tarlige).mp.<br>6 1 or 2 or 3 or 4 or 5<br>7 furosemide.mp. or Furosemide/<br>8 frusemide.mp.<br>9 bumetanide.mp. or Bumetanide/<br>10 (torsemide or torasemide).mp.<br>11 ethacrynic acid.mp. or Ethacrynic Acid/<br>12 "loop diuretic*".mp.<br>13 "Sodium Potassium Chloride Symporter Inhibitors"/<br>14 7 or 8 or 9 or 10 or 11 or 12 or 13<br>15 "prescribing cascade*".mp.<br>16 ("drug cascade*" or "medication cascade*").mp.<br>17 ("peripheral edema*" or "peripheral oedema*").mp.<br>18 ("drug-induced edema*" or "drug induced edema*").mp.<br>19 ("drug-induced oedema*" or "drug induced oedema*").mp.<br>20 ("sequence symmetry analysis" or "prescription sequence symmetry" or PSSA).mp.<br>21 Inappropriate Prescribing/<br>22 "prescription cascade*".mp.<br>23 Pharmacoepidemiology/ or pharmacoepidemiol*.mp.<br>24 15 or 16 or 17 or 18 or 19 or 20 or 21 or 22 or 23<br>25 6 and (14 or 24) |
| **Embase (Ovid)** | 23 April 2026 | 43 | 1 exp gabapentin/ or gabapentin.mp.<br>2 exp pregabalin/ or pregabalin.mp.<br>3 exp mirogabalin/ or mirogabalin.mp.<br>4 gabapentinoid*.mp.<br>5 (neurontin or lyrica or tarlige).mp.<br>6 1 or 2 or 3 or 4 or 5<br>7 "prescribing cascade*".mp.<br>8 "prescription cascade*".mp.<br>9 ("drug cascade*" or "medication cascade*").mp.<br>10 ("peripheral edema*" or "peripheral oedema*").ti.<br>11 ("drug-induced edema*" or "drug induced edema*").mp.<br>12 ("drug-induced oedema*" or "drug induced oedema*").mp.<br>13 ("sequence symmetry analysis" or "prescription sequence symmetry" or PSSA).mp.<br>14 7 or 8 or 9 or 10 or 11 or 12 or 13<br>15 6 and 14 |
| **PubMed** | 23 April 2026 | 241 | ("gabapentinoid*"[tiab] OR "gabapentin"[tiab] OR "pregabalin"[tiab] OR "mirogabalin"[tiab] OR "alpha-2-delta ligand*"[tiab] OR "neurontin"[tiab] OR "lyrica"[tiab] OR "tarlige"[tiab] OR "Gabapentin"[Mesh] OR "Pregabalin"[Mesh])<br>AND<br>("loop diuretic*"[tiab] OR "furosemide"[tiab] OR "frusemide"[tiab] OR "bumetanide"[tiab] OR "torsemide"[tiab] OR "torasemide"[tiab] OR "ethacrynic acid"[tiab] OR "prescribing cascade*"[tiab] OR "prescription cascade*"[tiab] OR "medication cascade*"[tiab] OR "peripheral edema"[tiab] OR "peripheral oedema"[tiab] OR "drug-induced edema"[tiab] OR "drug induced edema"[tiab] OR "sequence symmetry analysis"[tiab] OR "prescription sequence symmetry"[tiab] OR "PSSA"[tiab] OR "pharmacoepidemiol*"[tiab] OR "Sodium Potassium Chloride Symporter Inhibitors"[Mesh] OR "Furosemide"[Mesh] OR "Edema"[Mesh] OR "Inappropriate Prescribing"[Mesh]) |

| | | | |
|---|---|---|---|
| **Web of Science Core Collection** | 23 April 2026 | 233 | TS=("gabapentin*" OR "pregabalin*" OR "mirogabalin*" OR "gabapentinoid*" OR "alpha-2-delta ligand*" OR "neurontin" OR "lyrica" OR "tarlige")<br>AND<br>TS=("loop diuretic*" OR "furosemide" OR "frusemide" OR "bumetanide" OR "torsemide" OR "torasemide" OR "ethacrynic acid" OR "prescribing cascade*" OR "prescription cascade*" OR "medication cascade*" OR "peripheral edema*" OR "peripheral oedema*" OR "drug-induced edema*" OR "drug induced edema*" OR "drug-induced oedema*" OR "drug induced oedema*" OR "sequence symmetry analysis" OR "prescription sequence symmetry" OR "PSSA" OR "pharmacoepidemiol*" OR "inappropriate prescribing*") |

Figure S1. Preferred Reporting Items for Systematic Reviews and Meta-Analyses (PRISMA) flow chart.

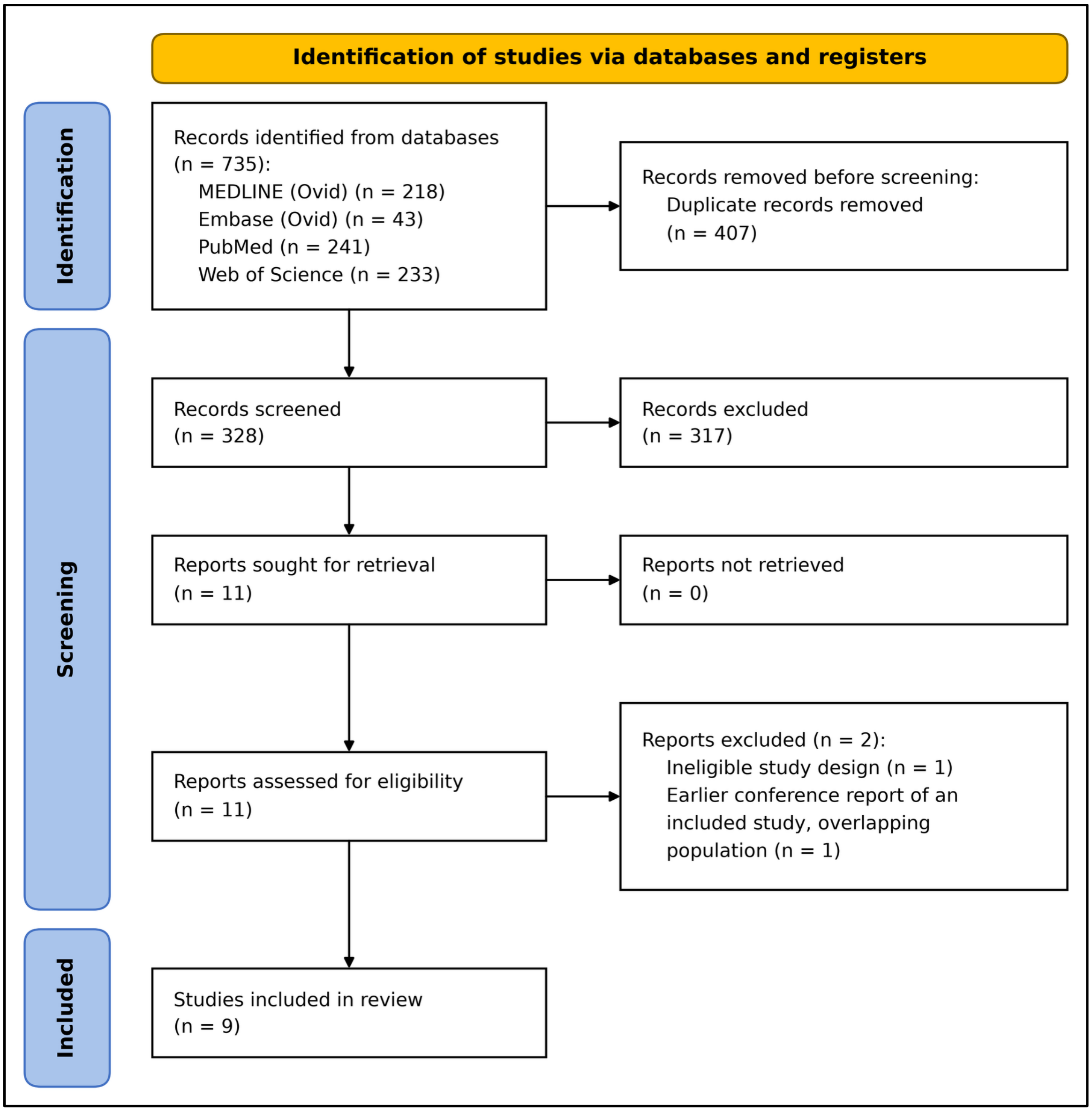

#### Supplementary Table S2. Risk estimates for the gabapentinoid–loop-diuretic prescribing cascade, overall and by dose, gabapentinoid agent, age, and time window, across the nine included studies.

| Study | Country | Design | Stratum | Comparison / level | Measure | Estimate (95% CI) | Window | Outcome |
|---|---|---|---|---|---|---|---|---|
| **Read 2021** | Canada | Cohort | Overall | Overall | Adjusted HR | 1.44 (1.23–1.70) | 90 d | Any diuretic |
| | | | Overall | Overall | Adjusted HR | 1.26 (1.11–1.43) | 180 d | Any diuretic |
| | | | By dose | High-dose gabapentinoid (time-varying) | Adjusted HR | 2.34 (1.17–4.67) | 180 d | Any diuretic |
| **Vouri 2022** | USA, Denmark | Sequence symmetry | Overall | Pooled (US + Denmark) | Adjusted SR | 1.33 (1.06–1.67) | 90 d | Loop diuretic |
| | | | Overall | USA | Adjusted SR | 1.19 (1.15–1.23) | 90 d | Loop diuretic |
| | | | Overall | Denmark | Adjusted SR | 1.50 (1.43–1.57) | 90 d | Loop diuretic |
| | | | By dose | < defined daily dose (US) | Adjusted SR | 1.17 (1.13–1.21) | 90 d | Loop diuretic |
| | | | By dose | ≥ defined daily dose (US) | Adjusted SR | 1.52 (1.32–1.74) | 90 d | Loop diuretic |
| | | | By drug | Gabapentin (pooled) | Adjusted SR | 1.31 (1.02–1.68) | 90 d | Loop diuretic |
| | | | By drug | Pregabalin (pooled) | Adjusted SR | 1.43 (1.26–1.62) | 90 d | Loop diuretic |
| | | | By age | < 65 y (pooled) | Adjusted SR | 1.34 (0.95–1.90) | 90 d | Loop diuretic |
| | | | By age | ≥ 65 y (pooled) | Adjusted SR | 1.38 (1.25–1.51) | 90 d | Loop diuretic |
| **Growdon 2024** | USA | Sequence symmetry | Overall | Overall | Adjusted SR | 1.23 (1.13–1.34) | 180 d | Loop diuretic |
| | | | By drug | Gabapentin vs pregabalin | Adjusted OR (cascade order) | 0.64 (0.40–1.02) | 180 d | Loop diuretic |
| | | | By age | ≥ 85 vs 66–74 y | Adjusted OR (cascade order) | 0.74 (0.56–0.96) | 180 d | Loop diuretic |
| | | | By age | 75–84 vs 66–74 y | Adjusted OR (cascade order) | 1.06 (0.90–1.25) | 180 d | Loop diuretic |
| | | | By polypharmacy | ≥ 10 vs 0–4 medications | Adjusted OR (cascade order) | 1.39 (1.07–1.82) | 180 d | Loop diuretic |
| **Growdon 2025** | USA | Chart review | Overall | Cascade cases (descriptive) | Downstream-harm frequency | 23.3% (28/120) | 60 d | Downstream harm |
| **Hayes 2026**[a] | USA | Cohort | Overall | Overall | Cumulative incidence | 4.2% (994/23,544) | 90 d | Loop diuretic |
| | | | By dose | Higher vs lower weekly dose (time-varying) | Risk ratio | 1.45 (7.2% vs 5.0%) | 13 wk | Loop diuretic |
| | | | By dose | High vs low baseline dose | Adjusted RR | 1.21 (0.99–1.48) | 90 d | Loop diuretic |
| | | | By drug | Pregabalin vs gabapentin | Adjusted RR | 1.15 (0.90–1.48) | 90 d | Loop diuretic |
| | | | By age | 71–75 vs 66–70 y | Adjusted RR | 1.14 (0.89–1.48) | 90 d | Loop diuretic |
| | | | By age | 76–80 vs 66–70 y | Adjusted RR | 1.21 (0.94–1.55) | 90 d | Loop diuretic |
| | | | By age | 81–85 vs 66–70 y | Adjusted RR | 1.16 (0.90–1.50) | 90 d | Loop diuretic |

| | | | | | | | | |
|---|---|---|---|---|---|---|---|---|
| | | | By age | 86–90 vs 66–70 y | Adjusted RR | 1.60 (1.25–2.04) | 90 d | Loop diuretic |
| | | | By age | ≥ 91 vs 66–70 y | Adjusted RR | 1.38 (1.06–1.79) | 90 d | Loop diuretic |
| **Omata 2024** | Japan | Sequence symmetry | Overall | Pregabalin + mirogabalin | Adjusted SR | 1.69 (1.56–1.83) | 90 d | Loop diuretic |
| | | | By drug | Mirogabalin | Adjusted SR | 2.23 (1.84–2.71) | 90 d | Loop diuretic |
| | | | By drug | Pregabalin | Adjusted SR | 1.59 (1.46–1.73) | 90 d | Loop diuretic |
| **Ito 2025** | Japan | Sequence symmetry | Overall | Pregabalin | Adjusted SR | 1.29 (1.16–1.43) | 90 d | Loop diuretic |
| | | | Overall | Pregabalin | Adjusted SR | 1.17 (1.09–1.25) | 360 d | Loop diuretic |
| | | | By dose | Starting dose < 150 mg/d | Adjusted SR | 1.19 (1.05–1.35) | 90 d | Loop diuretic |
| | | | By dose | Starting dose ≥ 150 mg/d | Adjusted SR | 1.53 (1.27–1.84) | 90 d | Loop diuretic |
| | | | By age | < 70 y | Adjusted SR | 1.33 (1.11–1.60) | 90 d | Loop diuretic |
| | | | By age | ≥ 70 y | Adjusted SR | 1.28 (1.13–1.45) | 90 d | Loop diuretic |
| **Henriksen 2020** | Denmark | Sequence symmetry | Overall | Overall | Sequence ratio | 1.56 (1.34–1.84) | 90 d | Loop diuretic |
| | | | By age | < 65 y (younger) | Sequence ratio | 1.74 (1.38–2.24) | 90 d | Loop diuretic |
| | | | By age | ≥ 65 y (older) | Sequence ratio | 1.43 (1.16–1.79) | 90 d | Loop diuretic |
| **Largeau 2022** | France | Cohort (matched) | Overall | Overall | Adjusted HR | 1.44 (1.20–1.73) | 120 d | Loop diuretic |
| | | | By time period | Days 1–15 | Adjusted HR | 0.96 (0.67–1.39) | 1–15 d | Loop diuretic |
| | | | By time period | Days 16–60 | Adjusted HR | 1.43 (1.12–1.84) | 16–60 d | Loop diuretic |
| | | | By time period | Days 61–120 | Adjusted HR | 2.01 (1.46–2.78) | 61–120 d | Loop diuretic |

*aHR/HR, (adjusted) hazard ratio; aSR/SR, (adjusted) sequence ratio; aRR/RR, (adjusted) risk ratio; aOR, adjusted odds ratio of cascade ordering; VA, Veterans Affairs.*

*[a] Hayes 2026 (full paper) and the Hayes 2025 ISPE conference abstract report the same study. Sequence-ratio (PSSA) and cohort (HR/RR/incidence) estimands are distinct and are not directly comparable. Growdon 2025 is a single-arm chart review; 23.3% is a descriptive harm frequency, not a cascade association. Largeau 2022 period-specific adjusted HRs are from the conference poster.*

### Figure S2. Random-effects meta-analysis of adjusted sequence ratios from full-text sequence symmetry analyses

This analysis examined whether quantitative pooling was defensible within the PSSA design. Eligible estimates were adjusted sequence ratios for loop diuretic initiation. We used the 90-day window where available (Vouri 2022, Omata 2024, Ito 2025) and otherwise the only reported window (Growdon 2024, 180 days). The United States and Danish estimates of Vouri 2022 were entered as separate data sources. Henriksen 2020 (conference abstract; Funen, Denmark, 2000–2016) was excluded from the main analysis because its population likely overlaps the Danish national data in Vouri 2022 (2005–2018), and was added in a sensitivity analysis. Between-study variance was estimated by restricted maximum likelihood, and confidence intervals were calculated with the Hartung–Knapp–Sidik–Jonkman method.

Main analysis (k = 5): pooled adjusted sequence ratio 1.37 (95% CI 1.14–1.64); $I^2 = 96.2\%$; Cochran's Q = 104.9 (4 df; $P < .001$). Given the very high heterogeneity, the pooled value is a descriptive summary of the sequence symmetry evidence.

Sensitivity analyses: adding Henriksen 2020, 1.39 (95% CI 1.20–1.62); 90-day estimates only, 1.40 (1.09–1.80); excluding Omata 2024 (pregabalin and mirogabalin only), 1.30 (1.10–1.54).

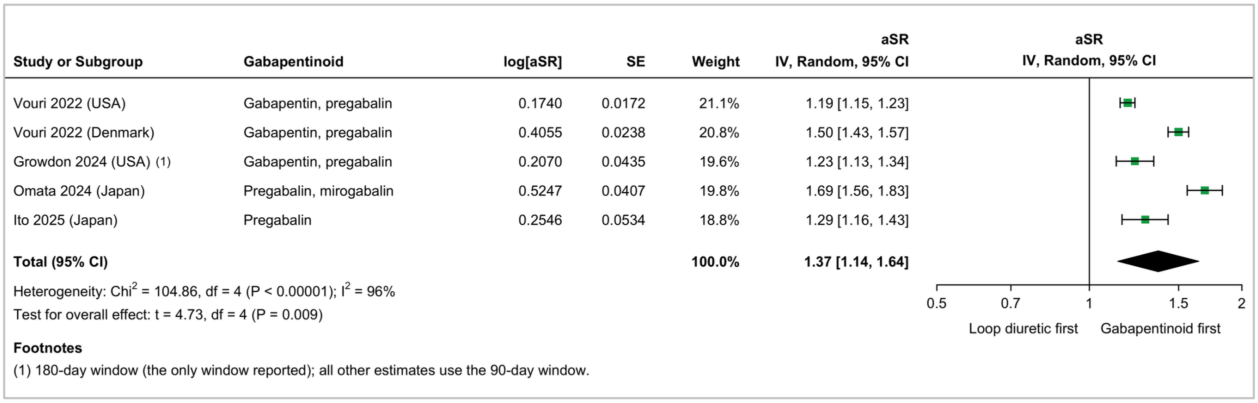


*aSR, adjusted sequence ratio; CI, confidence interval*

### Figure S3. Risk of bias of the seven studies with a comparison group (ROBINS-I), by domain

Risk-of-bias domains (ROBINS-I)

| | D1 | D2 | D3 | D4 | D5 | D6 | D7 | Overall |
|---|---|---|---|---|---|---|---|---|
| Growdon 2024 | – | X | + | + | + | + | – | X |
| Henriksen 2020 | – | ? | + | + | + | + | – | – |
| Ito 2025 | X | – | – | + | – | + | – | X |
| Largeau 2022 | X | – | + | – | – | + | – | X |
| Omata 2024 | X | + | + | + | + | + | + | X |
| Read 2021 | – | + | + | + | + | + | + | – |
| Vouri 2022 | – | – | + | + | + | + | + | – |
| Growdon 2025 | *Not assessed with ROBINS-I (no comparison group)* | | | | | | | |
| Hayes 2026 | *Not assessed with ROBINS-I (no comparison group)* | | | | | | | |

Domains:
D1: Bias due to confounding.
D2: Bias due to selection of participants.
D3: Bias in classification of interventions.
D4: Bias due to deviations from intended interventions.
D5: Bias due to missing data.
D6: Bias in measurement of outcomes.
D7: Bias in selection of the reported result.

Judgement
X Serious
– Moderate
\+ Low
? No information

Judgments refer to each study's principal result. For sequence symmetry analyses, domains were interpreted for the self-controlled design.

### Supplementary Table S3. Handling of concomitant edema-inducing medications and related time-varying factors in the included studies

| Study | Design | Concomitant edema-inducing drugs | Other relevant design features |
|---|---|---|---|
| **Read 2021** | Cohort | Prevalent NSAID users excluded; adjusted for use on the index date of drugs that may cause edema (including calcium channel blockers, NSAIDs, corticosteroids, and thiazolidinediones) | Heart failure, end-stage renal disease, and liver failure excluded; hospitalization in prior month excluded; adjusted for comorbidity and healthcare use |
| **Vouri 2022** | PSSA | Stratified by use of NSAIDs, calcium channel blockers, and oral corticosteroids in the 360 days before and after initiation; association present in all strata | Null-effect sequence ratio for secular trends; SNRI negative control; heart failure and CKD excluded |
| **Growdon 2024** | PSSA | Exposure to edema-associated drugs in the prior 6 months similar between prescribing sequences (57% vs 58%); not otherwise accounted for | Null-effect sequence ratio; heart failure, CKD, liver disease, and venous insufficiency excluded |
| **Growdon 2025** | Chart review | Not applicable (descriptive); non-gabapentinoid drug causes (mostly calcium channel blockers) were documented in 9.2% of patients (11/120) | Records restricted to documented edema; sample enriched for dementia |
| **Hayes 2026** | Cohort | Adjusted for baseline (prior 365 days) use of calcium channel blockers, corticosteroids, diuretics, and other co-medications | Heart failure and renal insufficiency excluded; adjusted for comorbidity and healthcare use |
| **Omata 2024** | PSSA | Not reported | Null-effect sequence ratio; heart failure and CKD retained and stratified |
| **Ito 2025** | PSSA | Not reported; time-dependent concomitant medication acknowledged as a limitation | Null-effect sequence ratio; duloxetine negative control |
| **Henriksen 2020[a]** | PSSA | Not reported | Heart failure and CKD excluded |
| **Largeau 2022[a]** | Cohort | Adjusted for baseline use of drugs that can cause edema | Matched on age, sex, hypertension, and month of initiation; heart failure and end-stage renal disease excluded |

*[a]Conference-only report. CKD, chronic kidney disease; NSAID, nonsteroidal anti-inflammatory drug; PSSA, prescription sequence symmetry analysis; SNRI, serotonin–norepinephrine reuptake inhibitor.*
